# Dutch participatory surveillance framework for evaluating evolutionary changes on SARS-CoV-2 affecting Rapid Diagnostic Test sensitivity in 2022 –2023

**DOI:** 10.1101/2024.09.10.24313404

**Authors:** Eva Kozanli, Wanda Han, Tara Smit, Jordy de Bakker, Mansoer Elahi, Ryanne Jaarsma, Gesa Carstens, Albert Jan van Hoek, Dirk Eggink

## Abstract

**Background:** Rapid Diagnostic Tests (RDTs) have been pivotal in the diagnostics for SARS-CoV-2 and policies surrounding self-isolation. When self-testing policies are in place a decreased sensitivity of the virus to RDTs can give benefits for viral spread. However, to monitor for reduced sensitivity of RDTs we rely on the collection of SARS-CoV-2 positive samples from RDT negative patients. *Infectieradar,* a national participatory surveillance that registers influenza-like symptoms is used as a framework to study false-negative RDT results due to emergence of new virus variants.

**Methods:** Participants report weekly on RDT use and symptoms linked to Acute Respiratory Illness (ARI). Each week, all RDT positive and a sample of 200 among RDT negative but symptomatic participants were invited to send in nose throat swabs (NTS). SARS-CoV-2 Ct-values are determined using RT-PCR on NTS samples for RDT positive and RDT (false) negative participants and compared. Sequencing is performed on all eligible samples for phylogenetic analysis of the SARS-CoV-2 nucleocapsid protein and the whole genome sequence. NTS samples of participants with discordant RT-PCR and RDT results are also analyzed using RDTs by professionals in the laboratory.

**Results:** Between October 2022 and October 2023, our study had 16,893 participants and we collected 1,757 self-test-positive/NTS PCR positive samples and 359 self-test-negative/NTS PCR positive samples (RDT-/PCR+). These participants were asked to take a SARS-CoV-2 RDT upon symptoms. Within SARS-CoV-2 PCR positive participants, we did not find characteristics that differ in SARS-CoV-2 RDT negative versus positive participants. There were no associations with specific changes in the N protein nor did our phylogenetic analysis show clustering of RDT negative samples.

**Conclusion:** Evaluating brand-specific RDT performance in Dutch population and false-negative RDT analyses, led to no evidence for SARS-CoV-2 evolution affecting RDT sensitivity. The participatory surveillance program *Infectieradar* is a powerful tool for our national surveillance on acute respiratory illnesses, as well as for research purposes. Since this framework offered both self-testing and the gold standard of PCR testing results.

## Background

Rapid Diagnostic Tests (RDTs) for SARS-CoV-2 played a significant role in controlling the COVID-19 pandemic and are still valuable for mitigating individual risk, surveillance and research. Since the start of the pandemic, self-isolation was based on SARS-CoV-2 testing, for which reverse transcriptase polymerase chain reaction (RT-PCR) was the diagnostic standard (1). When later SARS-CoV-2 self-tests, or RDTs, became available, they were distributed and sold widely, allowing convenient and less expensive self-testing at home, making downscaling of centralized test facilities possible(2). Although the pandemic is declared over, the sensitivity and specificity of RDTs are important to monitor routinely to ensure that they function appropriately even when new variants of the virus arrive.

Due to the wide use of RDTs and subsequent behavior when positive, such as isolation or other preventive action, the virus would benefit from remaining undetected in these tests, resulting in a false negative outcome. Furthermore, genetic drift and selective pressure on the virus, such as host-adaptation and immunity-escape could result in failing self-tests. By far most RDTs detect the nucleocapsid (N) protein of SARS-CoV-2. This N-protein is involved at multiple stages of the virus life cycle and is therefore abundant in SARS-CoV-2, making it a sensitive target for such RDTs. A potential risk is that mutations of the N-protein, or even changes in other regions of the genome, can weaken or abolish binding to antibodies used in the RDT and thereby affect its sensitivity(3, 4).

Routine monitoring of the sensitivity of RDTs is therefore important, both during a pandemic, and continuously afterwards. Low sensitivity may cause overlooking the virus, leading to a false sense of safety with a continuous spread of the virus, with subsequent implications for those infected. Monitoring this sensitivity requires confirming RDT negative tests by a gold standard, like RT-PCR.

It is known that sensitivity of RDTs is lowered in cases with low viral load, like in an asymptomatic population. Other contributions to lower sensitivity might be pointed out from false negative testing in symptomatic participants(5). As self-testing partially replaced the PCR-testing standard for detection of SARS-CoV-2, it is feasible that selective pressure drives viral strains to become more difficult to detect. Viruses continue to evolve over time and factors that seem to drive this are an important subject for study. The undetected SARS-CoV-2 in RDTs could be caused by biological and evolutionary changes in the virus, resulting in undetected circulation. As sensitivity of RDTs can depend on many factors, we attempt to focus on some of the viral characteristics to clarify part of it.

Earlier studies have performed epitope mapping to evaluate sites that are likely bound by antibodies and deep mutational scanning to look at less conserved sites of the N-protein, which could both inform us on potential risks of SARS-CoV-2 variant mutations for the efficacy of RDTs (6, 7). These sites can be monitored through sequence data to keep track of RDT sensitivity and monitoring is the responsibility of the RDT producers. Moreover, studies only found indirect evidence that SARS-CoV-2 variants affect the performance of a RDT (8, 9).

In our surveillance program, *Infectieradar* (10, 11), we have asked participants to perform a RDT when experiencing respiratory symptoms. Every week, participants received questionnaires and when they report a positive test result are asked to send in a self-sampled combined nose/throat swab (NTS) to confirm SARS-CoV-2 infection and determine viral load using RT-PCR, as well as determine the virus variant as part of the national SARS-CoV-2 genomic surveillance. A random subset of approximately 200 participants per week reporting a negative RDT after indicating respiratory symptoms is also asked to send in an NTS to identify circulating respiratory pathogens. Among this second group, SARS-CoV-2 is also sporadically detected, despite the reported negative RDT. This phenomenon is often missed in studies and raises questions about RDT sensitivity.

The set-up of *Infectieradar* allows us to investigate SARS-CoV-2 RDT sensitivity using PCR results from NTS samples. We provide a single brand of RDTs to participants, which makes self-testing accessible and uniform. As the focus of many studies have been mostly directed towards SARS-CoV-2 positive cases, our framework that monitored ARI might gather information on those cases that ought to be our blind spot: false negative RDT cases. We analyze these cases to evaluate the use of a participatory surveillance framework for ARI for monitoring and elaborate on its use in the future. Whole genome sequencing using an extensive set of RDT-negative samples and comparing those to RDT positive samples creates an opportunity to thoroughly study possible biological and evolutionary features of SARS-CoV-2 affecting RDT performance and to evaluate the current self-testing method.

## Methods

To monitor false negative RDTs per SARS-CoV-2 variant, we explored the RDT negatives per Ct-value and time in the pandemic, compared nucleocapsid mutations between positives and negatives, as well as phylogenetic clustering.

### Setup and sample collection

The setup of the *Infectieradar* study and participation of individuals is thoroughly described by Smit et al. (11). This study from the National Institute for Public Health and the Environment of the Netherlands (RIVM) is part of the *InfluenzaNet* network which studies acute respiratory illness (ARI) in various countries(12). Participants are followed through questionnaires filled out weekly concerning symptoms signaling acute infections, such as respiratory diseases as the common cold, COVID-19 or ARI but also gastrointestinal complaints and other infections (supplementary methods). *Infectieradar* recruited their participants among the general population, independently of healthcare seeking behavior.

Questionnaires are sent out weekly to 16.893 participants, in which they report their health and respiratory symptoms (supplementary methods). Participants were asked to do a SARS-CoV-2 RDT when they experience respiratory symptoms. As of October 2022, this study also contains a self-swab-component for nose throat swab (NTS), allowing laboratory RT-PCR based diagnostics for a broad panel of respiratory pathogens. A sampling kit was delivered to all participants upon inclusion, containing NTS, a vial containing Virus Transport Media (Xebios Diagnostics), rapid SARS-CoV-2 antigen RDTs (MP Biomedicals), appropriate return envelops suited for medical diagnostic materials and instructions including a link to the website with instruction video(10). Participants reporting a positive RDT were asked to send in an NTS. Out of the group that reported respiratory symptoms but a SARS-CoV-2-negative RDT, a weekly subset of 200 participants was randomly selected also sent an NTS. A ticketing-system was used to establish random sampling, which is explained in the paper of Smit et al. (11). Only participants that full-fill inclusion criteria are invited in this system. For declined invitations the system generates a new ticket to invite another participant using random sampling to ensure enough samples are collected per week. However, some participants with either a positive or negative RDT sent in their samples despite not being invited, their samples were included as well. Samples received at RIVM were stored at -80 degrees Celsius after performing molecular diagnostics.

### Diagnostics

#### RT-PCR

Firstly, SARS-CoV-2 viral load was determined for participants’ NTS samples through RT-PCR at the National Institute for Public Health and the Environment (RIVM), Bilthoven, The Netherlands as described (1). RNA extraction was performed using MagNApure 96 (MP96) with total nucleic acid kit small volume (Roche). 5 µL of the purified nucleic acid was used in TaqMan® Fast Virus 1-Step Master Mix (Thermo Fisher) in the Roche LC480 II thermal cycler. RT-PCR targeted the E-gene and RdRP-gene using specific primers and probe and EAV as controls. A Cycle threshold (Ct)-value lower than 40 was considered SARS-CoV-2 positive.

#### Sequencing

The RNA extract of each sample with a Ct-value below 32 was used in Amplicon-based sequencing, following the NanoPore protocol “PCR tiling of COVID-19 virus (Version: PTC_9096_v109_revE_06FEB2020)” derived from the ARTIC v3 protocol as described earlier(13, 14). Sequencing was performed on an Oxford Nanopore R9.4.1 flow cell. Sequences were only considered when there was less than 5% ambiguity.

### RDT negative and Ct-value: statistics

PCR results were analyzed separately for RDT+/PCR+ and RDT-/PCR+ group. Their frequencies and proportions were determined for each month to specify the contribution of the falsely negative group over time. Ct-values in each group are compared using Student’s unpaired T-test. Ct-values of the RDT-/PCR+ group over the time of this study were compared using linear regression. Visualizations of these analyses are made in R (v. 4.3.1).

### Nucleocapsid mutation analysis and phylogeny

SARS-CoV-2 whole genome were used to analyze presence of nucleocapsid substitutions. Sequences are then analyzed using NextClade (Web 2.14.1) for variant determination and to look at nucleocapsid substitutions(15). These mutations were used in an analysis to compare the occurrence of them in RDT+/PCR+ and RDT-/PCR+ participants. Visualizations are made using Python (v.3.10) and the seaborn package.

To compare sequences based on their RDT result, we aligned the curated sequences to a reference sequence (EPI ISL: 12954045) of a descendent omicron variant BA.1. We used this comparison to identify clustering between RDT negative samples. We downsampled sequences per ISO week and based on a positive RDT result to get an equal representation between RDT+/PCR+ and RDT-/PCR+ samples over time. The evaluated alignment was used to generate a phylogeny based on a maximum likelihood approach in IQtree (v. 1.6.12). GTR+F+R2 Model is used according to the Bayesian Information Criterion based on the embedded ModelFinder function(16, 17). The resulting tree is rooted in Figtree (v. 1.4.4) on our BA.1 reference sequence and the phylogeny is visualized using R (v. 4.3.1) and the ggplot2 package (18).

### RDT performance testing

RDT results are reported by each participant as described above. In case of discrepancies between the reported RDT result and RT-PCR results of the NTS, we tested the NTS sample (with Ct-values below 21) for reactivity in the RDT. RDT lysis buffer was mixed in a 1:1 ratio with NTS sample and 50 μl was added to the RDT cassette. After 15 minutes incubation at room temperature, results were scored by three independent assessors.

## Results

### Well-dispersed set of samples from participants reporting ARI symptoms

The study collected 528.350 questionnaires from 16.893 participants (55.1% female) that could be filled in every week between October 2022 and October 2023. In 99,044 questionnaires participants reported ARI symptoms. In this period, we received 7,274 NTS for follow-up with diagnostics, of which 75.1% of the samples were coupled to a reported RDT result. All sampled participants in this study were older than 16 years (median age = 53 years, 64.7% female).

### Diagnostic PCR showed low Ct-values in RDT positive and negative samples

Characteristics of the NTS samples are compared by coupling them to RDT results. We analyzed RDT and NTS results of 1,757 self-test-positive/NTS PCR positive samples (RDT+/PCR+) and 359 self-test-negative/NTS PCR positive samples (RDT-/PCR+) (4.3% false negative out of all RDT negative samples). The median time between the NTS and RDT was 0 days (RDT-/PCR+ median = 1 day). First, size and proportions of these groups were compared over the time of this study (see Figure 1). The share of RDT-/PCR+ within all SARS-CoV-2 PCR positive participants does not significantly differ over time of our study. This is indicative for a lack of increase in circulation of novel variants that possibly “escape” detection by RDTs.

**Figure 1.**
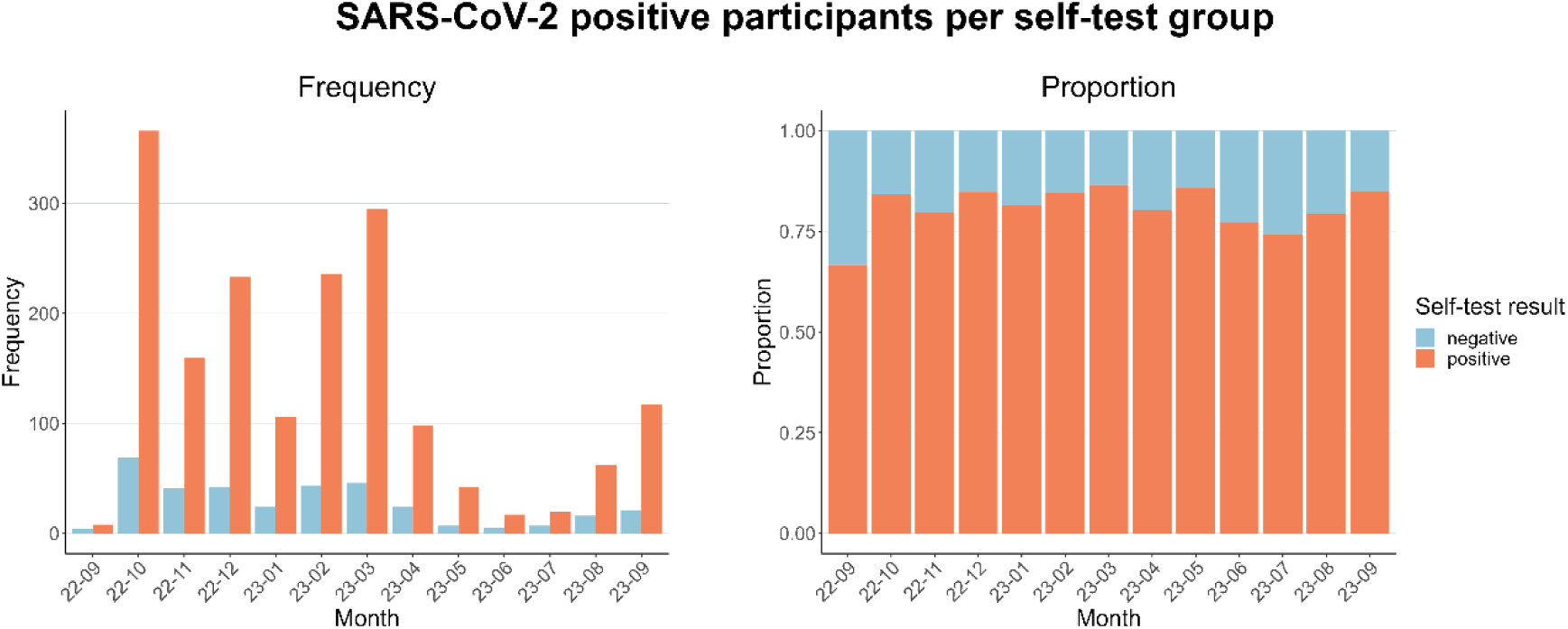
Frequency and proportion of participants with SARS-CoV-2 positive NTS results. This figure shows the frequency of SARS-CoV-2 positive NTS samples in each month during the study. Coloring is based on their RDT result.

To compare NTS samples from RDT+/PCR+ and RDT-/PCR+ participants, Figure 2 shows the distribution of Ct-values within the RDT-/PCR+ and RDT+/PCR+ group. The median Ct-value of the RDT-group was significantly higher than that of the RDT+ group which indicates a generally lower viral load for the RDT-group. Yet, we also find low Ct-values in the NTS results for a significant number of participants in the RDT negative group, ruling out low viral load in participants as sole determinant for failing self-tests. This first and lowest quantile (highest viral load) for the negative group is of interest, as failing methodology (such as suboptimal sampling) is less likely to occur for these NTS samples and viral properties could contribute to the lack of detection in RDT.

**Figure 2.**
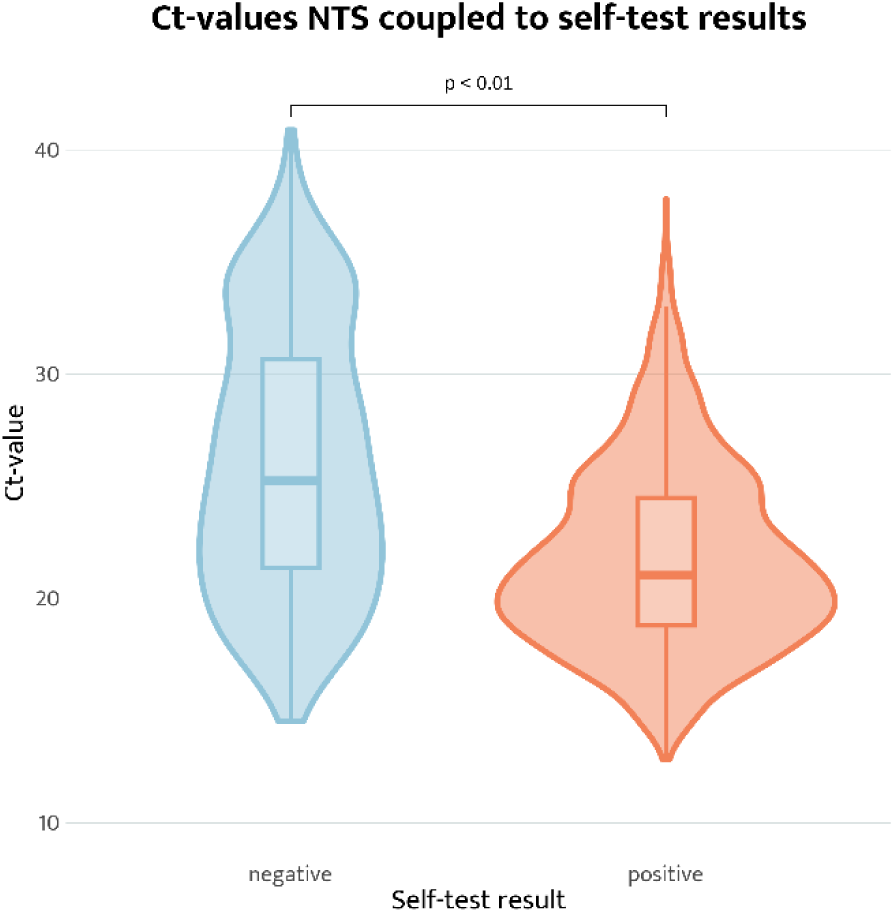
Violin plot of Ct-values of NTS samples with positive or negative RDT. Figure shows NTS samples of RDT+/PCR+ and RDT-/PCR+ group (n= 1757, n=359). Ct-value of the RDT-/PCR+ group is significantly higher than that of the RDT+ group. Low Ct-values (<20) are found in both groups.

A first indication for changes in the virus itself is shown in Figure 3. When analyzing the Ct-values from the NTS over time for the RDT negative group, no correlation is observed (P = 0.809), again indicative of absence of increase of circulation of novel SARS-CoV-2 variants that are not detected by RDTs.

**Figure 3.**
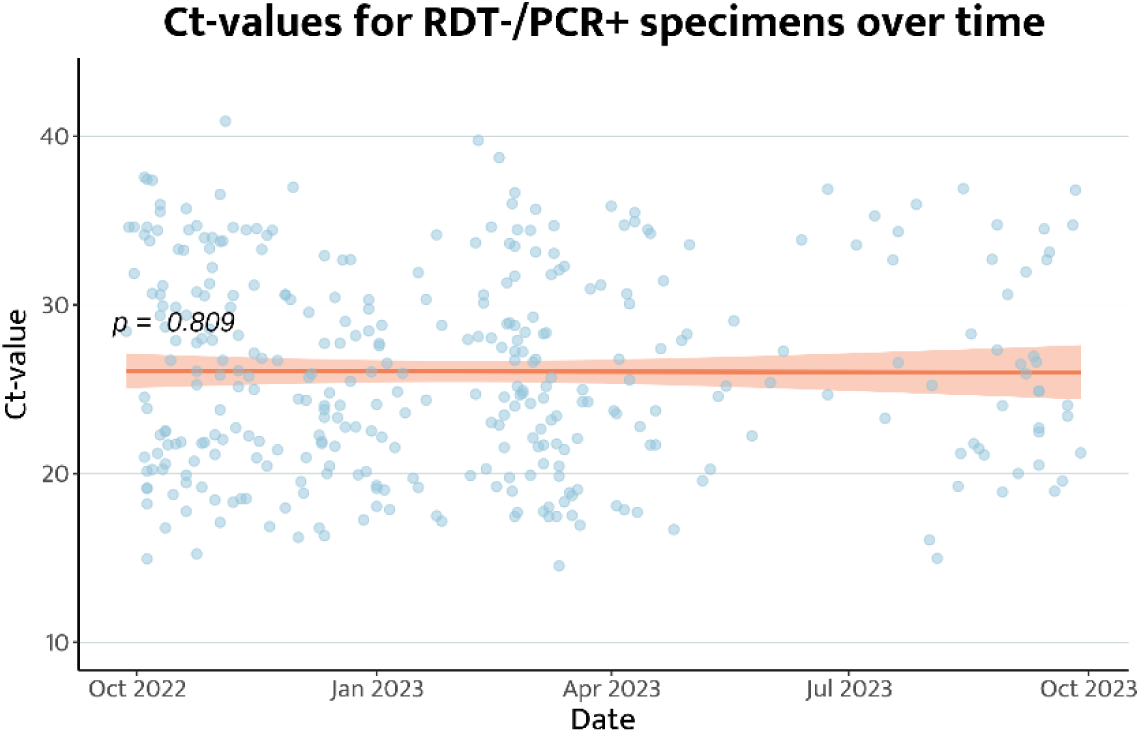
Ct-values of NTS samples for RDT-/PCR+ participants over time of the study. Linear regression of the Ct-value for negatively self-tested participants shows no correlation between Ct-value and time of sampling (n= 359, P = 0.809).

### Nucleocapsid substitutions are similar in RDT positive and negative groups

SARS-CoV-2 positive NTS samples were sequenced using NanoPore to identify variants. This allows characterization of weekly trends that can be compared to nationwide sequencing programs. The comparison (Figure 4) shows that we have obtained a set of samples that are representative of the general population. Firstly, we compared the nucleocapsid substitutions in our sequences, to identify presence or over-representation of specific changes in the N protein that could explain the lack of detection of N protein by the RDT. The figure shows a unique substitution in the RDT-/PCR+ group for N: A182S. This substitution occurred in a higher frequency for this group (p < 0.05, Chi-Squared test), but still occurred in only 2 sequences. We have not found this substitution to be linked to failed detection by RDTs in literature. These sequences both date from the start of our study and we did not find an increase in this mutation over time. The higher frequency of the deletion in GER30-32 and substitution S33F are not found to be significant (Chi-Squared Test). These alternative frequencies are not proportional to the complete set of sequences and are therefore not expected to cause evolution towards RDT evasion at large scale. Based on prevalence analyses of substitutions within the RDT+/PCR+ and RDT-/PCR+ group, we did not find substitutions that were disproportionate in either one of these groups, that could explain failed detection by the RDT (Figure 5).

**Figure 4.**
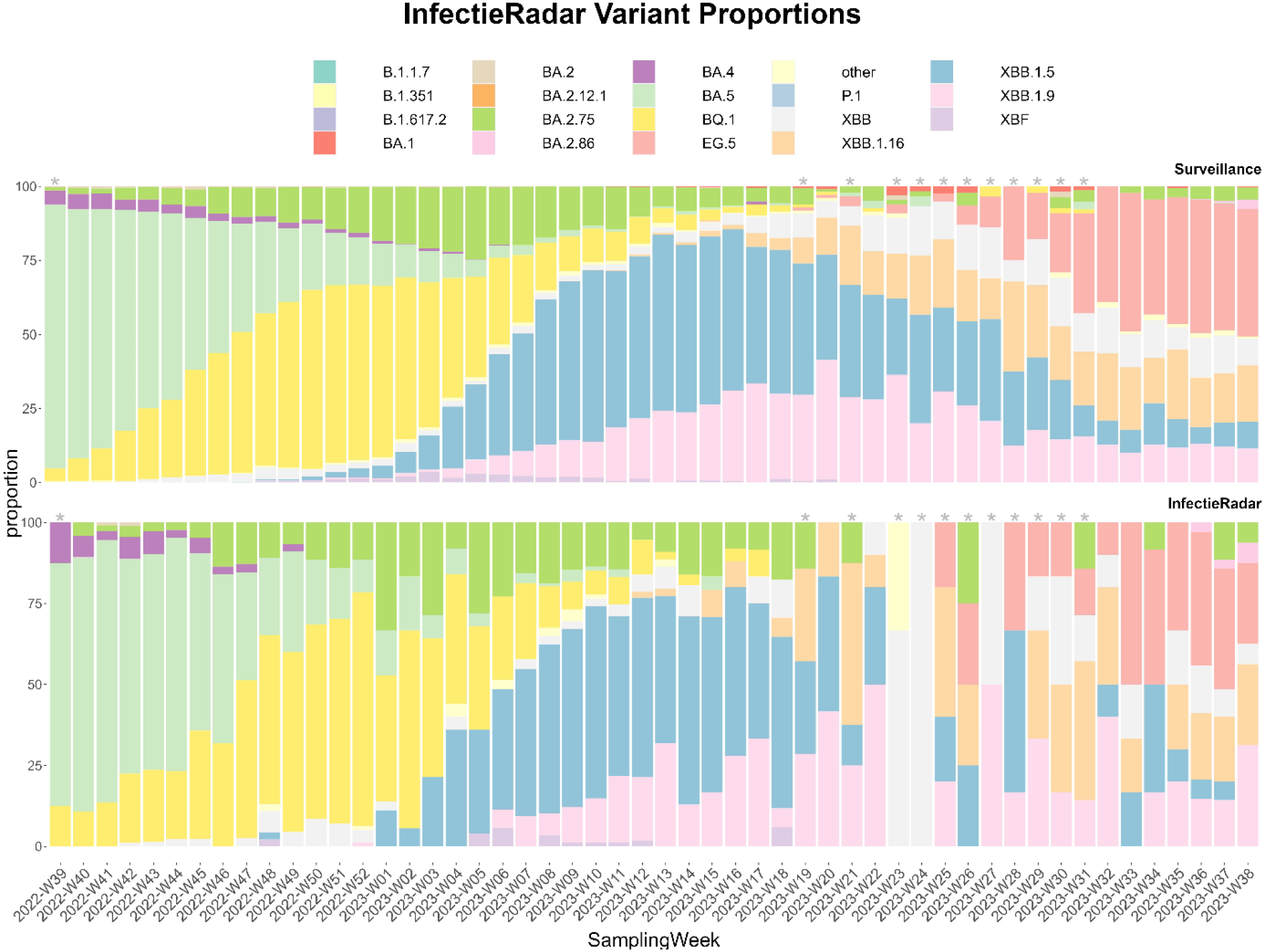
*Infectieradar* Variant Proportions. The figure shows the variant detection for each of the *Infectieradar* samples compared to the national surveillance variant detection. These show similar patterns for each week. Most weeks have a representative set of sequences. The weeks that have a low number of sequences (< 10), are indicated with the asterisk (*).

**Figure 5.**
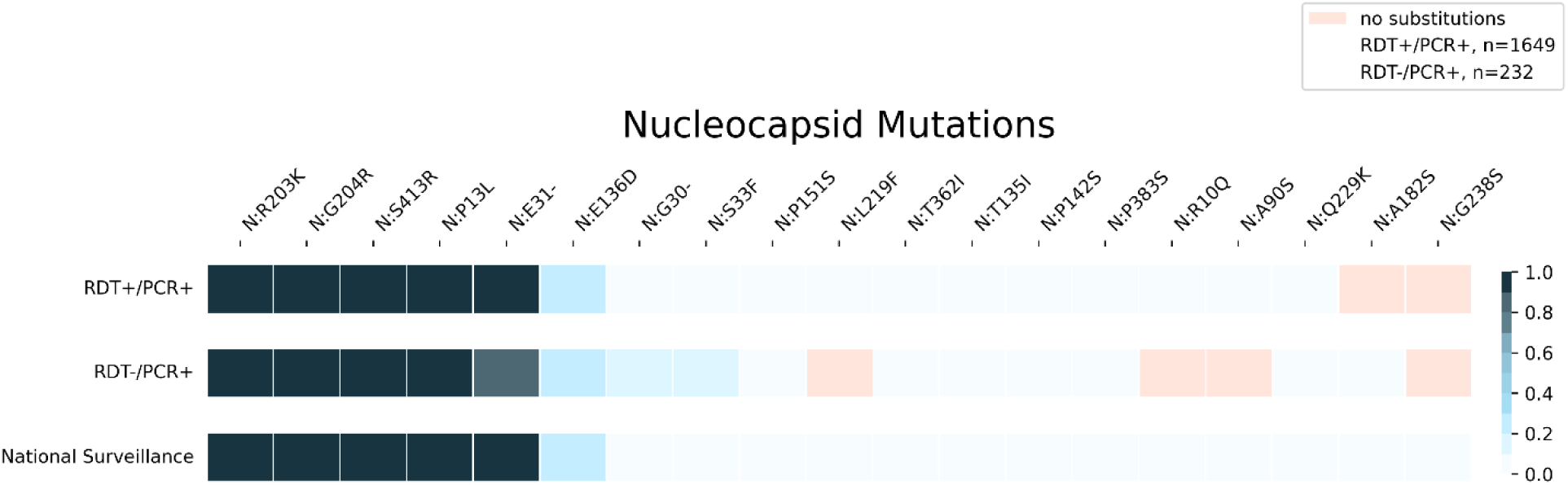
Heatmap of Nucleocapsid mutations based on *Next Clade* typing. Figure shows minimal differences between RDT negative and positive samples and national surveillance.

### Phylogenetic analysis showed no clustering of negative RDT samples

To analyze possible grouping of (genetic) SARS-CoV-2 variants detected in the samples from the RDT negative group, indicating a combination of genetic changes in N and other genomic regions responsible for diminished detection by RDT, a phylogenetic tree is generated to look at genetic similarity of RDT negative virus isolates. Sequence analyses show wide diversity of Omicron subvariants present during the period of this study. As mentioned above, these sequences reflect variants circulating as measured in the National Genomic SARS-CoV-2 surveillance of the Netherlands(19). The phylogeny in Figure 6 shows a downsampled enriched for RDT negative samples. The sequences are rooted on a previous BA.1 sequence (EPI ISL: 12954045). Red tips indicate RDT+/PCR+ isolates and blue stands for RDT-/PCR+ participants. The tree does not show clustering of RDT-/PCR+ isolates but shows that sequences are widely dispersed over the tree and co-appear with genomes obtained from SARS-CoV-2 viruses obtained from RDT+ samples.

**Figure 6.**
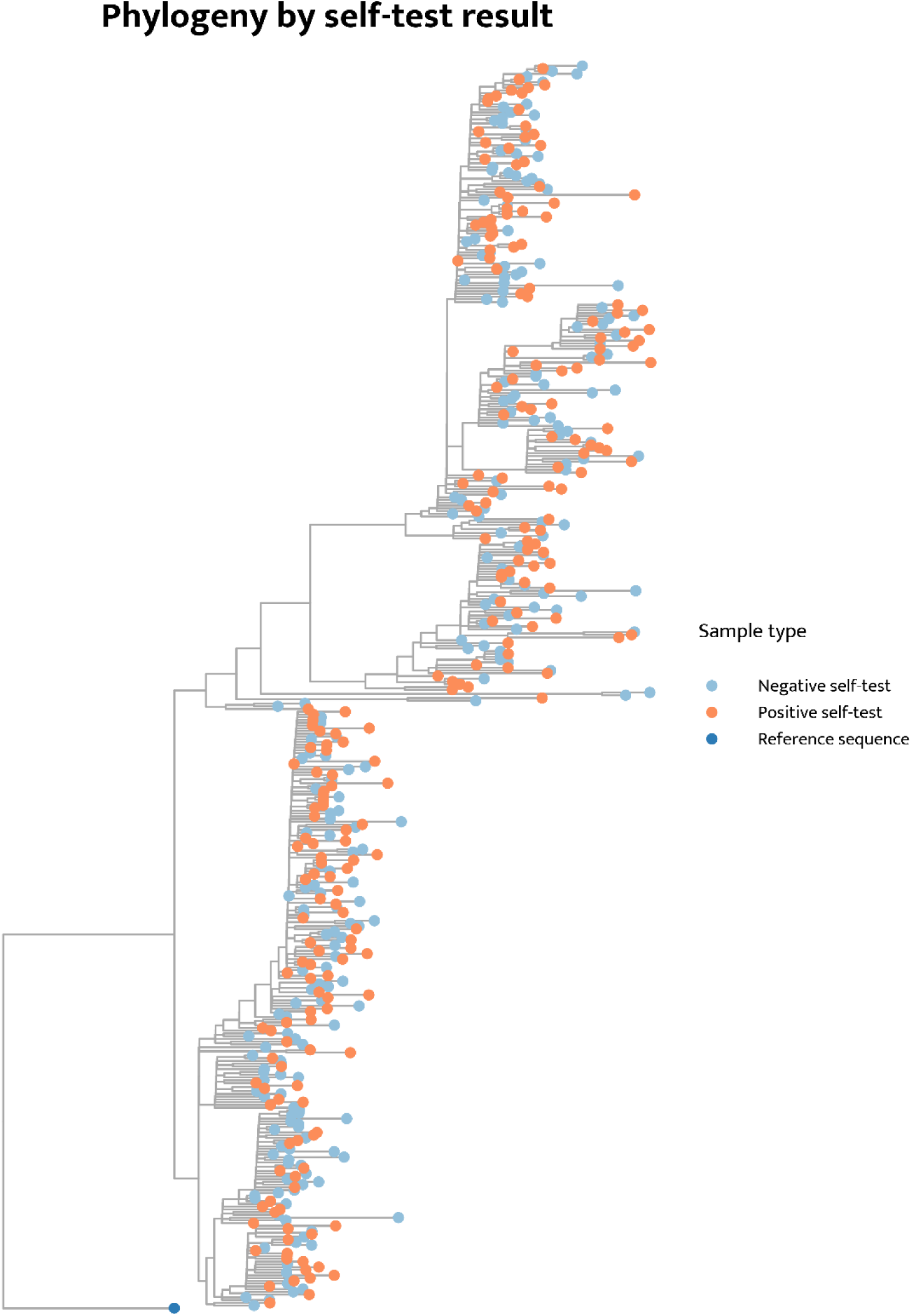
Phylogeny based on RDT results. This figure shows the phylogenetic tree of SARS-CoV-2 isolates. Red indicates RDT+/PCR+ samples and blue shows RDT-/PCR+ samples. Sequences from RDT negative samples do not cluster but are spread through the different branches of the tree.

### Laboratory-based RDT duplicate generated positive test results

To account for the effect of methodology on falsely negative self-tests, we reproduced RDTs using the sent left-over NTS material of each RDT negative participant that had an NTS with a Ct-value below 21 (n = 75). Samples from the RDT-/PCR+ group showing the lowest Ct-values (highest viral load) were selected. All samples tested did result in a positive self-test, as assessed by three assessors, confirming our PCR and sequencing approaches. The NPS sample containing the unique N: A182S mutation also resulted in a positive RDT in the laboratory.

## Discussion

Real life, on-going validation of performance of SARS-CoV-2 RDTs remains important for their use in decreasing viral spread by self-isolation. We show that routine collection of paired NTS and RDT results in a participatory surveillance system providing a way to monitor the functioning of RDTs and possible related genetic features. By analyzing 7,274 NTS samples of symptomatic participants with ARI, we were able to compare PCR and sequence results in depth for a SARS-CoV-2 RDT positive and negative group. Even in times of high genetic diversity of the virus and policy implemented which advised self-testing as ground for self-isolation, we found no indication of altered RDTs sensitivity due to evolutionary changes in SARS-CoV-2. We did not find an increase of the false negative contribution to our samples. Also, we did not see selection of substitutions in the nucleocapsid, nor did we find clustering when analyzing the entire genome profile in negative RDT isolates. This leads us to conclude that these RDTs can properly detect currently circulating variants and that there is no evidence that genetic drift affected SARS-CoV-2 self-test performance.

We found that low viral load in participants contributes to false negative self-testing, shown by higher Ct-values in the paired sample analyzed by PCR in the RDT negative group which indicates challenges for sensitivity of the tests. However, we would describe viral load in participants to have a mainly supporting role, since low Ct-values are also observed in the paired PCR result for a considerable number of RDT negative participants. Our suggestion is that self-sampling methodology of RDT users contributes to false negative RDT results in which case suboptimal self-sampling, RDT sensitivity compared to PCR and difficulties in reading a weak signal on the RDT may play a role. The PCR+ NTS samples of participants (with Ct-values lower than 21) that reported a negative RDT result, were found RDT positive in the laboratory, providing some evidence that discordant results between PCR and RDT results are possibly due to suboptimal sampling of the sample used for self-testing and/or execution of the RDT by participants.

There are some notes to be taken when interpreting our conclusion. Sampling for self-testing by RDTs and NTS for PCR testing might not have been taken simultaneously for each participant, but we observed that for most cases, they were taken within the same day or the next day. Tropism of the virus also plays a role in this comparison since for RDTs only the nose was sampled, while for PCR nose and throat were sampled, which can affect viral load in the tested sample. Like most studies, we struggle with a bias in participation which we hoped to have reduced with the accessibility of sampling and questionnaires. Although samples were not stratified for participant specific characteristics, our ticketing-system ensured more randomness for sample selection. Our evaluation of this system is positive, since it yielded a representative number of samples for most weeks. Some participants sent in a sample without being invited, which was possible because each participant had a new sampling set at hand, already before inviting to ensure quick sampling in our study.

During this study, the use of RDTs was encouraged by providing all participants with tests and resupplying when necessary. This led to the use of RDTs from a single brand, of which extensive in-house validation has taken place beforehand. Quality tests comparing various SARS-CoV-2 variants have been done to test performance of this test and put this into context with other RDTs (20). The conclusion of our study can therefore be used as reference to pursue studying RDT sensitivity for the evolving SARS-CoV-2 virus, but not as a conclusion for the sensitivity of all available RDTs on the market. When compared to national surveillance, our dataset appears to be representative. Our diagnostic PCR is robust as it is extensively checked and updated based on national surveillance data, leading to consistency in our measured Ct-values. The above qualities provide a strong argument for the *Infectieradar* program, as a scalable surveillance framework that provides a nearly real-time dataset. Due to the high and constant engagement of participants in questionnaires, we reached them shortly after symptom onset. The large sample size of this study led to a representative group of false negatives and their NPS samples helped us verify their results in the laboratory through the NPS. Participant-based sampling makes *Infectieradar* accessible for participants and cost-efficient.

An understanding of the sensitivity and specificity of RDTs is essential when interpreting surveillance data. New variants and developing immunity levels in the population seemed to lead to more asymptomatic cases. Updating these results is therefore necessary if used in future scenarios. Since *Infectieradar* is a broad surveillance program with sampling based on ARI symptoms it facilitates monitoring multiple respiratory viruses at once. This overarching initiative can result in more cost-effective monitoring. Not only does this study about monitoring RDT performance inform future policymaking, but it also provides opportunities for research and evaluation by utilizing the programs longitudinal and scalable character. This way*, Infectieradar* can contribute to pandemic preparedness too.

## Conclusion

In conclusion, our analyses show no evolutionary changes of SARS-CoV-2 that contributed to false-negative results by RDTs. The study emphasizes that our participatory surveillance system, with active sampling based on symptoms, contributes to both the national disease surveillance as well as the monitoring and evaluation of RDTs.

## Supporting information

Supplementary Material: questionnaires

## List of abbreviations

ARI: acute respiratory infection
COVID-19: coronavirus disease 2019
Ct-value: cycle threshold value
NTS: nose throat sample
RDT: Rapid diagnostic test
RIVM: Dutch National Institute for Public Health and the Environment (Rijksinstituut voor Volksgezondheid en Milieu)
RT-PCR: reverse transcriptase polymerase chain reaction
SARS-CoV-2: Severe acute respiratory syndrome coronavirus 2

## Ethics approval and consent to participate

The samples were collected as part of *Infectieradar*. The protocol for *Infectieradar* was approved by the Medical Ethics Review Committee Utrecht (reference number: WAG/avd/20/008757; protocol 20-131) was obtained given the nature of data collection. All participants provided informed consent upon registration, which included agreement to the privacy statement and described the processing of personal data and research results, website security measures taken, and how to file a complaint. Participants were eligible to withdraw from the study at any time. Individuals had to be 16 years or over to be able to participate.

## Consent for publication

Not applicable.

## Availability of data and material

The processed data required to reproduce the findings of this study are available from the corresponding author upon reasonable request. Due to confidentiality concerns, access to the raw data may be restricted.

## Authors’ contributions

WH, TS, AJdH, DE were responsible for the conceptualization and design of the study and participated in the acquisition of data; DE, JdB, ME, RJ coordinated and performed laboratory analyses; EK, WH, AJdH, DE, GC were responsible for data analyses, interpretation of the analyses and verified the underlying data; EK wrote the manuscript with review and editing put from all authors. DE, AJdH, WH were responsible for resources, supervision, project administration and funding acquisition. All authors had full access to all the data and reviewed and approved the final version of the manuscript.

## Acknowledgements

We want to thank all the participants of *Infectieradar* for their active and continuous participation. Without their contribution this project wouldn’t have been a success. Furthermore, we want to thank the *Infectieradar*-team at RIVM for all their efforts to keep the website up and running, handle the logistics, perform routine analysis and answer any questions of participants.

