## Supplementary Material: questionnaires for "Dutch participatory surveillance framework for evaluating evolutionary changes on SARS-CoV-2 affecting Rapid Diagnostic Test sensitivity in 2022 –2023"

### Questionnaire Intake Infectieradar

General remark: this questionnaire is originally published in Dutch but has been translated to English for international scientific publication purposes. Therefore, grammar, style or specific terms might differ from British or American English.

The intake survey focuses on some background and demographic information.

What is your sex at birth?

- Male
- Female
- Other

What is your date of birth (year and month)?

- Output: date –> feedback: [output] years old

What are the first four digits of your home postal code (the part before the space)?

- Output: 4 digits

What is your height (in cm)?

- Output: height
- I would rather not share this information/ I don’t know

What is your weight (in kg)?

- Output: weight
- I would rather not share this information/ I don’t know

What is your main activity?

- Paid employment, full time
- Paid employment, part time
- Self-employed (businessperson, farmer, tradesman, etc.)
- Attending daycare/school/college/university
- Homemaker
- Unemployed
- Long-term sick leave or parental leave
- Retired
- Other

What are the first 4 digits of the place where you spend most of your time, other than your home? For example your (volunteer) work or school/university?

- Output: 4 digits
- I don’t know
- Not applicable

On an average week how often do you go to your (volunteer) work or school/university?

- Less than 1 day
- 2 days
- 3 days
- 4 days
- 5 days
- More than 5 days

Which of the following descriptions most closely matches with your main occupation?

- I work in health care
- I have a job in which I am in close contact with others (for example hairdresser)
- I work in child care / education (primary school, secondary school, MBO, HBO, University)
- I work in a hotel, restaurant of café (horeca)
- I work in a shop (supermarket or other kind of shop)
- I work in public transport (for train, bus, metro, taxi)
- I am a knowledge worker (manager, researcher, accountant)
- I have an administrative role (administration, financial assistant, receptionist)
- I work with my hands (builder, technic, production)
- Different, not mentioned above

if working in education or childcare:

Where are you employed in education or childcare?

- I work in a nursery/childcare
- I work in primary education, or BSO
- I work in secondary education (VMBO/havo/VWO)
- I work at the MBO
- I work at the HBO or WO
- Other

if working in health care:

Where are you employed within health care?

- I work in a hospital
- I work in a nursing home
- I work in a care home
- I work in mental health
- I work at a GP practice
- I provide other kind of care (for example physiotherapy)
- Other, not mentioned above

What is the highest level of formal education/qualification that you have? (levels are indicated according to the Dutch system)

- I have no formal qualification

- MAVO. VMBO or equivalent
- HAVO, VWO, MBO or equivalent
- HBO or WO Bachelors Degree (BA, BSc) or equivalent
- Higher Degree or equivalent (e.g. Masters Degree, PhD, Medical Doctorate)
- I would rather not share this information/ I don't know

Except people you meet on public transports, do you have contact with any of the following during the course of a typical day (so without covid measures)?

- More than 10 children or teenagers
- More than 10 people aged over 65
- Groups of people (more than 10 individuals at any one time)
- None of the above

How many household members do you have?

- None
- 1
- 2
- 3
- 4
- 5
- 6
- 7
- 8
- More than 8

If household members indicated:

What is (are) your household member(s) age group(s)?

- 0-4 years old
- 5-12 years old
- 13-18 years old
- 19-44 years old
- 45-64 years old
- 65+

How many of the children in your household go to school or day-care?

- None
- 1
- 2
- 3
- 4
- 5
- More than 5

On a normal day, how much time do you spend on public transport? (Bus, train, metro etc.)

- No time at all
- 0 – 30 minutes
- 30 minutes to 1.5 hours
- 1.5 – 4 hours
- More than 4 hours

Have you already been infected with the coronavirus?

- No, I don't think I have been infected with the coronavirus
- I don't know
- Yes perhaps. I had/ have symptoms that look like it
- Yes, I do think so. I had symptoms, just as people around me
- Yes, I am pretty sure, I and people around my had symptoms, and one or more of those people tested positive
- Yes, I know for sure, as I tested positive and I had/have symptoms
- Yes, I know for sure, I tested positive, however I did not have any symptoms

Do you take regular medication for any of the following medical conditions?

- No
- Asthma
- Diabetes
- Chronic lung disorder besides asthma e.g. COPD, emphysema, or other disorders that affect your breathing
- Heart
- An immunocompromising condition (e.g. splenectomy, organ transplant, acquired immune deficiency, cancer treatment)
- I would rather not share this information/ I don't know

Are you currently pregnant?

- Yes
- No
- I would rather not share this information/ I don't know

If yes:

Which trimester of the pregnancy are you in?

- First trimester (week 1 to 12)
- Second trimester (week 13 to 28)
- Third trimester (week 29 to delivery)
- I would rather not share this information/ I don't know

Do you smoke tobacco?

- No
- Yes, occasionally

- Yes, daily, fewer than 10 times a day
- Yes, daily, 10 times or more times a day
- Yes, only e-sigarets
- I would rather not share this information/ I don’t know

Do you have one of the following allergies that can cause respiratory symptoms?

- Hay fever
- Allergy against house dust mites
- Allergy against domestic animals or pets
- Other allergies that cause respiratory symptoms (e.g. sneezing, runny eyes)
- I do not have an allergy that causes respiratory symptoms

Do you follow a special diet?

- No special diet
- Vegetarian
- Veganism
- Low-calorie
- Other

Do you have pets at home?

- No
- Yes, one or more dogs
- Yes, one or more cats
- Yes, one or more birds
- Yes, one or more other animals

How often do you experience common colds or flu-like diseases?

- Never
- Once or twice a year
- Between 3 and 5 times a year
- Between 6 and 10 times a year
- More than 10 times a year
- I would rather not share this information/ I don’t know

Did you receive a flu vaccine during the last autumn/winter season (2022/2023)?

- Yes
- No
- I would rather not share this information/ I don’t know

Are you planning to receive a flu vaccine this autumn/winter season (2023/2024)?

- Yes, I am planning to
- Yes, I got one

- No
- I don’t know (yet)

If they did receive a vaccine:

When were you vaccinated against flu in the season (2023/2024)?

- Output: date

If they did receive a vaccine:

What are your reasons for getting a seasonal influenza vaccination this year (2023/2024)?

- I belong to a risk group (e.g, pregnant, over 65, underlying health condition, etc)
- Vaccination decreases my risk of getting influenza
- Vaccination decreases the risk of spreading influenza to others
- My doctor recommended it
- It was recommended in my workplace/school
- The vaccine was readily available and vaccine administration was convenient
- The vaccine was free (no cost)
- I don't want to miss work/school
- I always get the vaccine
- I try to protect myself against infections, because of the circulation of the coronavirus
- Other reason(s)

If they did  not receive a vaccine:

What are your reasons for NOT getting a seasonal influenza vaccination in season (2023/2024)?

- I don't belong to a risk group
- It is better to build your own natural immunity against influenza
- I doubt that the influenza vaccine is effective
- Influenza is a minor illness
- I don't think I am likely to get influenza
- I believe that influenza vaccine can cause influenza
- I am worried that the vaccine is not safe or will cause illness or other adverse events
- I don't like having vaccinations
- The vaccine is not readily available to me
- The vaccine is not free of charge
- No particular reason
- Although my doctor recommend a vaccine, I do not get one
- Other reason(s)

Did you get a pneumococcal vaccine last winter season (2022/2023)?

- Yes
- No
- I would rather not share this information/ I don’t know

Did you receive a corona vaccine during the last autumn/winter season (2022/2023)?

- Yes
- No
- I would rather not share this information/ I don't know

Are you planning to receive a corona vaccine this autumn/winter season (2023/2024)?

- Yes, I am planning to
- Yes, I have had one
- No
- I don't know (yet)

If they did get a vaccine:

When were you vaccinated against corona in the season (2023/2024)?

- Output: date
- I would rather not share this information/ I don't know

If they did get a vaccine:

What are your reasons for getting a corona vaccination this year (2023/2024)?

- I belong to a risk group (e.g, pregnant, over 65, underlying health condition, etc)
- Vaccination decreases my risk of getting coronavirus
- Vaccination decreases the risk of spreading coronavirus to others
- My doctor recommended it
- It was recommended in my workplace/school
- The vaccine was readily available and vaccine administration was convenient
- The vaccine was free (no cost)
- I don't want to miss work/school
- I always get the vaccine
- I try to protect myself against infections, because of the circulation of the coronavirus
- Other reason(s)

If they did not get a vaccine:

What are your reasons for NOT getting a corona vaccination in the autumn/winter season (2023/2024)?

- I don't belong to a risk group
- It is better to build your own natural immunity against coronavirus
- I doubt that the coronavirus vaccine is effective
- Corona is a minor illness
- I don't think I am likely to get coronavirus
- I believe that corona vaccine can cause coronavirus
- I am worried that the vaccine is not safe or will cause illness or other adverse events
- I don't like having vaccinations
- The vaccine is not readily available to me

- The vaccine is not free of charge
- No particular reason
- Although my doctor recommend a vaccine, I do not get one
- Other reason(s)

On a scale from 0 to 100, how good or bad was your health last week?

- Output: number between 0 and 100

### Questionnaire symptoms (weekly) Infectieradar

Did you receive a corona test result since the last survey? (positive or negative)?

- No, I did not receive a test result
- Yes, I received the result of a self-test
- Yes, I received the result of a throat/nose swab (PCR)
- Yes, I received the result of a blood test (antibody test)

The following questions are about the self-test:

- How many self-tests did you take since the last questionnaire?
- 1
- 2
- 3
- More than 3

The following questions are about the self-test:

What was your self-test result?

- Positive, evidence for infection with coronavirus
- Negative, NO evidence for infection with coronavirus
- I prefer not to say

The following questions are about the self-test:

What was the date of the self-test? Please guess if you can't remember the date exactly.

- Output: Choose date

The following questions are about the nose/throat swab:

What was the date of the nose/throat swab? Please guess if you can't remember the date exactly.

- Output: Choose date

Where did you get yourself tested?

- Hospital or general practitioner
- At a commercial company ( own initiative)
- At a commercial company ( via employer)

- Abroad
- I do not know
- Self-test (Infectieradar self-swab study)

The following questions are about the nose/throat swab:

What was your nose/throat swab test result?

- Positive, evidence for infection with coronavirus
- Negative, NO evidence for infection with coronavirus
- I would rather not share this information/ I don’t know

The following questions are about the antibody test:

On which day was the blood taken for the corona test? Please guess if you can't remember the date exactly.

- Output: choose date

What was your blood test result?

- Positive, evidence for infection with coronavirus
- Negative, NO evidence for infection with coronavirus
- I would rather not share this information/ I don’t know

Please choose if you had any of the following symptoms since your last survey.

Did you have any general symptoms such as

- No symptoms
- Fever
- Chills
- Runny or blocked nose
- Sneezing
- Sore throat
- Cough
- Shortness of breath
- Headache
- Muscle/ joint pain
- Chest pain
- Feeling tired or exhausted (malaise)
- Loss of appetite
- Colored sputum/phlegm
- Watery, bloodshot eyes
- nausea
- vomiting

- diarrhea ( at least three times a day)
- stomach ache
- loss of smell
- Loss of taste
- Nose bleed
- Rash
- Other, namely [fill in]

When no symptoms are indicated, this is where the questionnaire ends.

When any symptoms are indicated it continues:

On a scale from 0 to 100, how good or bad was your health last week because of the symptoms?

The scale goes from 0 to 100, 100 means the best health status you can imagine and 0 means the worst health status you can imagine.

- Output: scale from 1 to 100

When symptoms were reported in last questionnaire as well:

On your last visit, you reported that you were still ill. Are the symptoms you report today part of the same bout of illness?

- Yes
- No
- I don't know/can't remember

When symptoms were not reported in last questionnaire:

When did the first symptoms appear? Please give as accurate an estimate as possible.

- Output: date

When did your symptoms end?

- Output: date
- I am still ill

Did your symptoms develop suddenly over a few hours?

- Yes
- No
- I don't know/can't remember

When fever was indicated:

When did your fever begin?

- Output: date
- I don't know/can't remember

When fever was indicated: Did your fever develop suddenly over a few hours?

- Yes
- No
- I don't know/can't remember

When fever was indicated: Did you take your temperature?

- Yes
- No
- I do not know

When yes was indicated:

What was your highest temperature measured?

- Below 37.0 °C
- 37.0 – 37.4 °C
- 37.5 – 37.9 °C
- 38.0 – 38.9 °C
- 39.0 – 39.9 °C
- 40.0 °C or more
- I don't know/can't remember

For all symptoms:

Do you also know where you may have contracted the infection?

- I don’t know
- Yes, abroad
- Yes, at home by a family member or roommate
- Yes, at home by someone who I was visiting
- Yes, at work
- Yes, at school
- Yes, during an activity in my spare time
- Yes, but not in a place mentioned above (e.g. people in the public transport or in shops)
- I don't want to indicate
- Not applicable, the symptoms are not related to an infection

Do you also know the gender and/or age range of the possible source or sources?

- No, I don't know
- Yes
- I would rather not share this information

If the gender/age are known:

What is the gender?

- Male
- Female
- Not clear, multiple possible sources
- I would rather not share this information

Which age group? (multiple answers possible)

- 0 – 3 years
- 4 - 6 years
- 7 – 12 years
- 13 – 18 years
- 19 – 29 years
- 30 – 39 years
- 40 – 49 years
- 50 – 59 years
- 60 – 69 years
- 70 – 79 years
- 80 – 89 years
- 90+ years
- I would rather not share this information/ I don’t know

Because of your symptoms, did you VISIT (see face to face) any medical services?

- No, I did not seek medical help
- Yes, GP or GP's practice nurse
- Yes, hospital accident & emergency department / out of hours service
- Yes, hospital admission
- Yes, other medical services
- Not yet, but I have an appointment scheduled

Did you visit the GP practice for your consult with the GP?

- No, the consult happened by phone or video-connection (video consult)
- Yes, I went to the GP practice to consult the GP

How soon after your symptoms appeared did you first VISIT a medical service?

- Same day
- 1 day
- 2 days
- 3 days
- 4 days
- 5 days
- 6 days
- 7 days
- 8 days
- 9 days
- 10 days
- 11 days
- 12 days
- 13 days
- 14 days
- More than 14 days
- I don’t know/ can’t remember

Did you take medication for these symptoms? Select all options that apply.

- No medication
- Yes, pain killers (e.g. paracetamol, lemsip, ibuprofen, aspirin, calpol, etc)
- Yes, cough medication (e.g. expectorants)
- Yes, hay fever medication
- Yes, nasal spray
- Yes, antivirals (Tamiflu or Relenza)
- Yes, antibiotics
- Yes, homeopathy
- Yes, alternative medicine ( essential oils, phytotherapy etc.)
- Yes, other medicine
- I don’t know/ can’t remember

If antivirals are indicated:

How long after the beginning of your symptoms did you start taking antiviral medication?

- Same day (within 24 hours)
- 1 day
- 2 days
- 3 days
- 4 days
- 5 days
- 6 days

- 7 days
- More than 7 days
- I don’t know/ can’t remember

Did you change your daily routine because of your illness?

- No
- Yes, but I did not take time off work/school
- Yes, I took time off work/school
- Not applicable, I don't have work/school

If time taken off from work/school:

Are you still off work/school?

- Yes
- No
- Other (e.g. I wouldn’t usually be at work/school today anyway)

If time taken off from work/school:

How long have you been off work/school? (only count the days you had to be there)

- 1 day
- 2 days
- 3 days
- 4 days
- 5 days
- 6 to 10 days
- 11 to 15 days
- More than 15 days

What do you think is the main reason causing your symptoms?

- Flu or flu-like illness
- Common cold
- Allergy/hay fever
- Asthma
- Gastroenteritis/gastric flu
- Coronavirus or corona-like illness
- Because I got vaccinated
- Other
- I don’t know
